# Community-level characteristics of COVID-19 vaccine hesitancy in England: A nationwide cross-sectional study

**DOI:** 10.1101/2022.03.15.22272362

**Authors:** Bucyibaruta Georges, Blangiardo Marta, Konstantinoudis Garyfallos

**Affiliations:** MRC Centre for Environment and Health, Department of Epidemiology and Biostatistics, School of Public Health, Imperial College London, London, UK

**Keywords:** COVID-19, Spatial modelling, Vaccine inequalities, Community-level characteristics

## Abstract

One year after the start of the COVID-19 vaccination programme in England, more than 43 million people older than 12 years old had received at least a first dose. Nevertheless, geographical differences persist, and vaccine hesitancy is still a major public health concern; understanding its determinants is crucial to managing the COVID-19 pandemic and preparing for future ones. In this cross-sectional population-based study we used cumulative data on the first dose of vaccine received by 01-01-2022 at Middle Super Output Area level in England. We used Bayesian hierarchical spatial models and investigated if the geographical differences in vaccination uptake can be explained by a range of community-level characteristics covering socio-demographics, political view, COVID-19 health risk awareness and targeting of high risk groups and accessibility. Deprivation is the covariate most strongly associated with vaccine uptake (Odds Ratio 0.55, 95%CI 0.54-0.57; most versus least deprived areas). The most ethnically diverse areas have a 38% (95%CI 36-40%) lower odds of vaccine uptake compared with those least diverse. Areas with the highest proportion of population between 12 and 24 years old had lower odds of vaccination (0.87, 95%CI 0.85-0.89). Finally increase in vaccine accessibility is associated with higher COVID-19 uptake (OR 1.07, 95%CI 1.03-1.12). Our results suggest that one year after the start of the vaccination programme, there is still evidence of inequalities in uptake, affecting particularly minorities and marginalised groups. Strategies including prioritising active outreach across communities and removing practical barriers and factors that make vaccines less accessible are needed to level up the differences.

## 1 Introduction

Mass vaccination has been an essential tool to fight the global COVID-19 pandemic. The National Health Service (NHS) in England began the vaccination programme in early December 2020 [1, 2] and around 43 million people have received at least the first dose of the vaccine by the end of 2021 [3]. Nevertheless, the uptake varies across population subgroups; vaccine inequalities continue to be a major public health concern and understanding their determinants is crucial to managing the COVID-19 pandemic and to prepare for future ones [4].

Several studies have examined the determinants of COVID-19 vaccine hesitancy across the globe [5, 6]. A study in 138 countries reported income disparity to be a main cause of vaccine inequity between middle-income and high-income countries [7]. Higher distrustful attitudes towards vaccination were reported amongst individuals from ethnic minority backgrounds, with low education, low annual income and lack of awareness of COVID-19 health risks in Qatar, Israel, New Zealand and USA [8–11]. Similarly, in the UK there has been evidence of lower intention to vaccinate in participants from Black and South Asian communities compared with the White population [12–14]. Additionally, less affluent areas have been reporting lower uptake, after accounting for individual level demographics and health conditions [15–17]. Some studies assessed a link between political believes and COVID-19 vaccine uptake. For example in the USA, people who identified themselves as Republicans or voted for the Republican party in the 2020 presidential election were less likely to get the vaccine [18, 19]. Inequality in vaccination-site accessibility was also identified as one of the challenges that recipients faced when attending vaccination appointments[20].

Certain methodological aspects of previous studies limit the generalisability of the results. Several earlier works are surveys, experiencing potential issues of statistical power and lack of population representativeness [5, 6, 18, 21–24]. Additionally, scientific evidence to date focuses on determinants of vaccine hesitancy prior or at the early stages of the mass vaccination campaigns [12–14, 24, 25]. As vaccination-related interventions (e.g. COVID-19 pass in some countries) and scientific evidence about vaccine efficacy have changed over time, people’s attitude towards COVID-19 vaccination is expected to change [4, 26]. Only two population-based studies covered a longer time period [10, 15]. The first analysed data at county level in the US until 29 July 2021, nevertheless the geographical resolution available was low, aggravating the ecological bias (group level associations that do not reflect individual ones [27]). The second is a register-based study in England which covered the period from the start of the mass vaccination up to 15 June 2021. However, due to the age prioritisation in the vaccine delivery by the NHS, its target population included only people aged 40 and over.

This is the first nationwide cross-sectional investigation of vaccine uptake in England during the entire 2021, covering the population aged 12 and older. We estimate the vaccine coverage and evaluate its determinants at a high geographical resolution. We extracted the reported cumulative data of COVID-19 vaccine as of the 1st of January 2022 in each area. To overcome the selection bias due to the age-based prioritisation programme of the government [28] we focus on the first dose of the vaccine. We consider community-level characteristics to cover socio-demographics, awareness of COVID-19 health risks and targeting of high risk groups, political views and vaccine accessibility. We account for spatial autocorrelation across neighboring areas and estimate the degree of geographical variability in vaccine uptake explained by the community-level characteristics considered.

## 2 Methods

### 2.1 Study area and variable of interest

We retrieved COVID-19 vaccination data at the Middle Layer Super Output Areas (MSOA) from the UK government dashboard [29]. MSOA is an administrative geography characterised by an average population of 7500 residents (varying between 5000 and 15000)[30]. We considered the 2011 geography, which comprises of 6791 MSOAs in England. To measure the vaccination uptake we considered the number of people aged 12 years and older in each MSOA, who had received at least one dose of COVID-19 vaccine since the beginning of the vaccination programme until the 1st January 2022. Total population for the same age group in each MSOA was used as the denominator.

### 2.2 Community-level characteristics

To examine the determinants of vaccine uptake at high spatial resolution, we considered covariates related with socio-demographics, political-opinion, COVID-19 mortality during 2020, mental and physical chronic health conditions as well as vaccine accessibility (see Table 1). To characterise the socio-demographic profile of each area we used percentage of Black and Minority ethnic (BME), index of multiple deprivation (IMD), percentage of 12-24 years old and percentage of over 65 years old in each MSOA. We classified each MSOA based on its level of urbanicity (Predominantly Urban (PU), Urban with Significant Rural (UR), and Predominantly Rural (PR)). To characterise the political-opinion of an area, we used the percentage who voted to leave the EU at the 2016 referendum and the results from the 2019 General election. To describe the awareness of the COVID-19 health risks and the targeting of high risk groups, we included the COVID-19 mortality rates during 2020 [31], covering the pre-vaccination campaign period, as well as the prevalence of asthma, high blood pressure, diabetes, and depression. The COVID-19 vaccine accessibility was estimated based on the distance between vaccination sites and MSOA population weighted centroids (see section S1 and Figure S1 in Supplementary Material for more details).

**Table 1.** Community-level characteristics considered in the analysis

| Variable | Description | Source | Spatial resolution |
| --- | --- | --- | --- |
| Black and minority ethnic (BME) | proportion of BME population | Public Health England [33] | Middle Super Output Area |
| Index of multiple deprivation (IMD) | measure of deprivation across multiple domains | Ministry of Housing, Communities and Local Government [34] | Lower Super Output Area |
| Young age population | proportion of 12-24 years old eligible for vaccine | Office for National Statistics [35] | Middle Super Output Area |
| Old age population | percentage of over 65 years old eligible for vaccine | Office for National Statistics [35] | Middle Super Output Area |
| COVID-19 awareness | rates of COVID-19 mortality | Office for National Statistics [36] | Local Authority District |
| Urbanicity | Rural and urban classification | Office for National Statistics[37] | Middle Super Output Area |
| General election | votes for Labour and Conservatives | House of Commons Library [38] | Constituency |
| EU referendum | votes to leave the EU | House of Commons Library [39] | Constituency |
| Pre-existing health conditions | asthma, blood pressure, diabetes, depression | National Health Service [40] | MSOA |
| Vaccine accessibility | estimated based on distance between vaccination sites and MSOA population weighted centroids | National Health Service [41]<br>Office for National Statistics[42] | Middle Super Output Area |

Information on the data sources and spatial resolution are presented in Table 1. All variables are included in the model in quintiles, except for urbanicity that has three categories. The shapefiles of England and MSOAs boundaries were obtained from the UK-data-service website [32].

### 2.3 Statistical analysis

We specified a hierarchical Bayesian spatial model to investigate the association of COVID-19 vaccine uptake and community-level characteristics. We considered *y*_*i*_ to be the number of people who have received the first dose of COVID-19 vaccine and *n*_*i*_ the number of eligible people to receive the vaccine for each MSOA (*i* = 1, …, 6791). We assumed a Binomial distribution for the number of vaccinations and modeled the proportion *p*_*i*_ as follows:

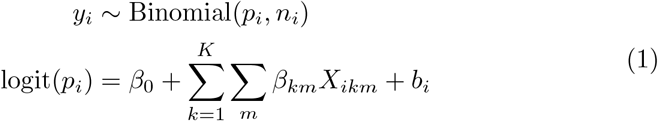

where *β*_0_ is the overall vaccination uptake across England, *X*_*ikm*_ is the dummy variable for the *k*^*th*^(*k* = 1, …, *K*) covariate in the *m*^*th*^ category (*m* = 2, …, 5, except for urbanicity where *m* = 2, 3) and ***β*** are the corresponding effects. Additionally, *b*_*i*_ represents the weighted average of a spatially structured and unstructured random effect, so that the model borrows strength from the other areas across the entire study region, as well as from the neighbouring ones [43, 44]. The random effects are modeled using a re-parametrisation of the Besag-York-Molli’e conditional autoregressive prior distribution [45]:

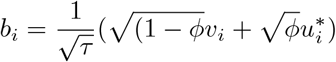

where *v*_*i*_ ∼ *N* (0, 1) accounts for overdispersion and 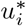 is a scaled spatially structured component. The hyperparameter *ϕ* measures the proportion of the marginal variance explained by the structured spatial effect, with values close to 0 implying that the majority of the observed variation comes from the unstructured (overdispersion) component (and values close to 1 the opposite). The hyperparameter *τ* is the precision of the random effect (1/variance). More details about the prior specification are included in Section S2 of Supplementary Material.

We report maps of the posterior median of vaccination uptake and highlight the patterns across the four largest cities. The effects of the covariates are reported as median odds ratios (OR) and 95% credible intervals (95%CI). We show and contrast the profiles based on the community-level characteristics included in the analysis for the areas in the lowest and highest quintile of vaccination uptake. We also report maps of posterior probability that the area-level odds ratio of vaccination is lower than the national average. This highlights the residual spatial variability, over and above that explained by the community-level characteristic profiles. Finally, we estimate the median and 95%CI for the proportion of total variance explained by the covariates. All analysis were conducted using the R statistical software and the INLA package [46]. Code and data to reproduce the results are available at https://github.com/Georges3/COVID_19-VaccineUptake.

## 3 Results

We report the results of the fully adjusted model, while those from the univariate models are showed in Supplementary Material (Figure S7, Table S5-S7 in Supplementary material). We estimated a national posterior mean of vaccine uptake of 81.1% (95%CI 80.2%-81.9%), varying from 37.6% (95%CI 36.6%-38.6%) in Leeds city in Yorkshire to 93.9% (95%CI 93.4%-94.5%) in Northumberland, a rural area in the North East. We observe a large geographical discrepancies in the vaccination coverage, with the lowest values in the large urban centres (Figure 1, left). Focusing on the four most populated cities, a high degree of heterogeneity can be seen, with lower vaccination uptake in the city centres (Figure 1, right). Spatial variability is also visible in the maps of the covariates, which are included in Supplementary Material (Figures S2 to S4).

**Fig. 1.**
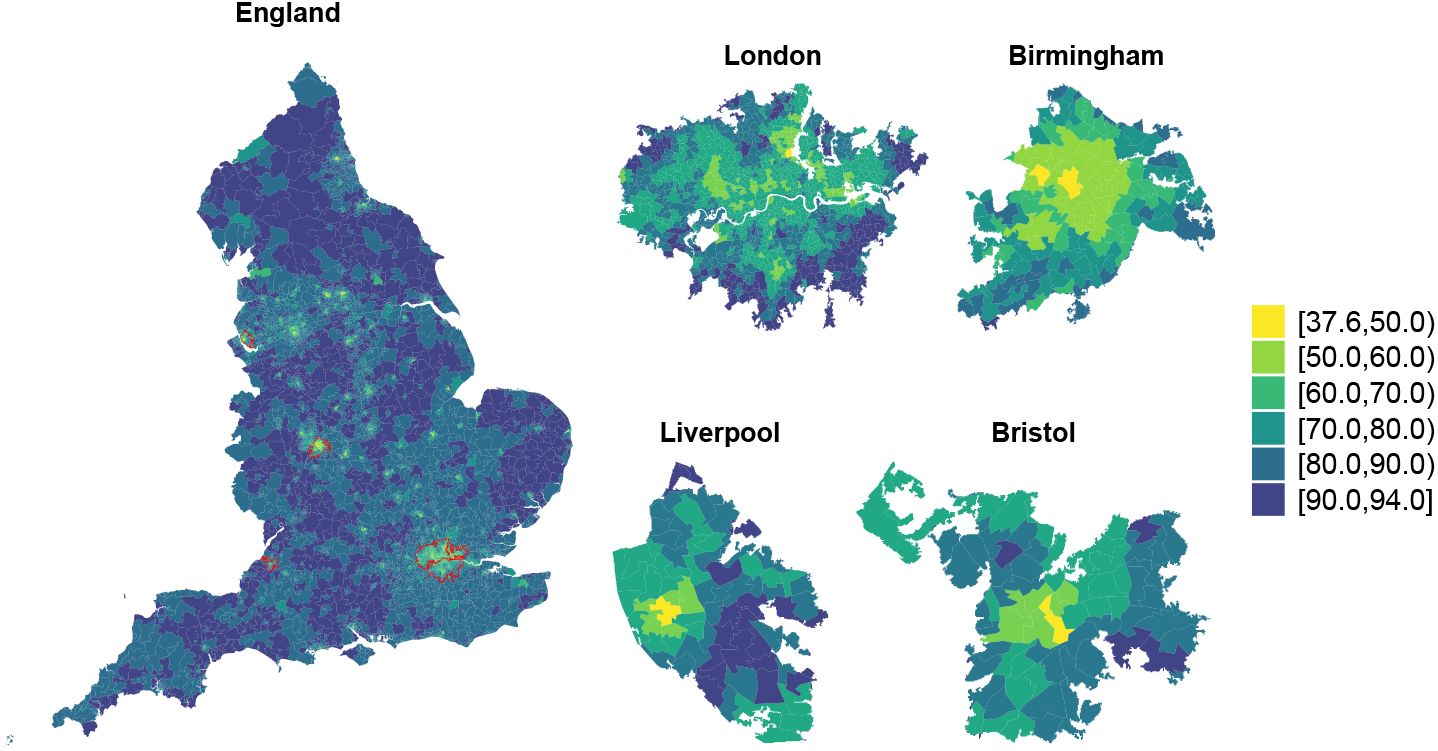
Posterior mean of vaccine uptake up until 01-01-2022 in each Middle Layer Super Output Area in England (left) and for the most populated cities accounting for a total 11 212 813 population [47] (right).

In Figure 2 we characterise the areas with the lowest estimates of vaccine uptake (in the first quintile). We visualise (a) their covariate profiles and (b) the posterior 95%CI of their vaccination uptake. Generally the least compliant areas share some characteristics: they tend to be more deprived and located in urban settings; the have high proportions of young residents and some of the highest proportions of non-White population. Based on the last general election, they are more inclined to vote for the Labour party. They also have the lowest prevalence of chronic conditions such as asthma, high blood pressure and depression. In contrast, the areas in the highest quintile of vaccine coverage are characterised by an older population, higher prevalence of asthma and blood pressure and vote mainly for the Conservative party (Figure S5 Supplementary Material).

**Fig. 2.**
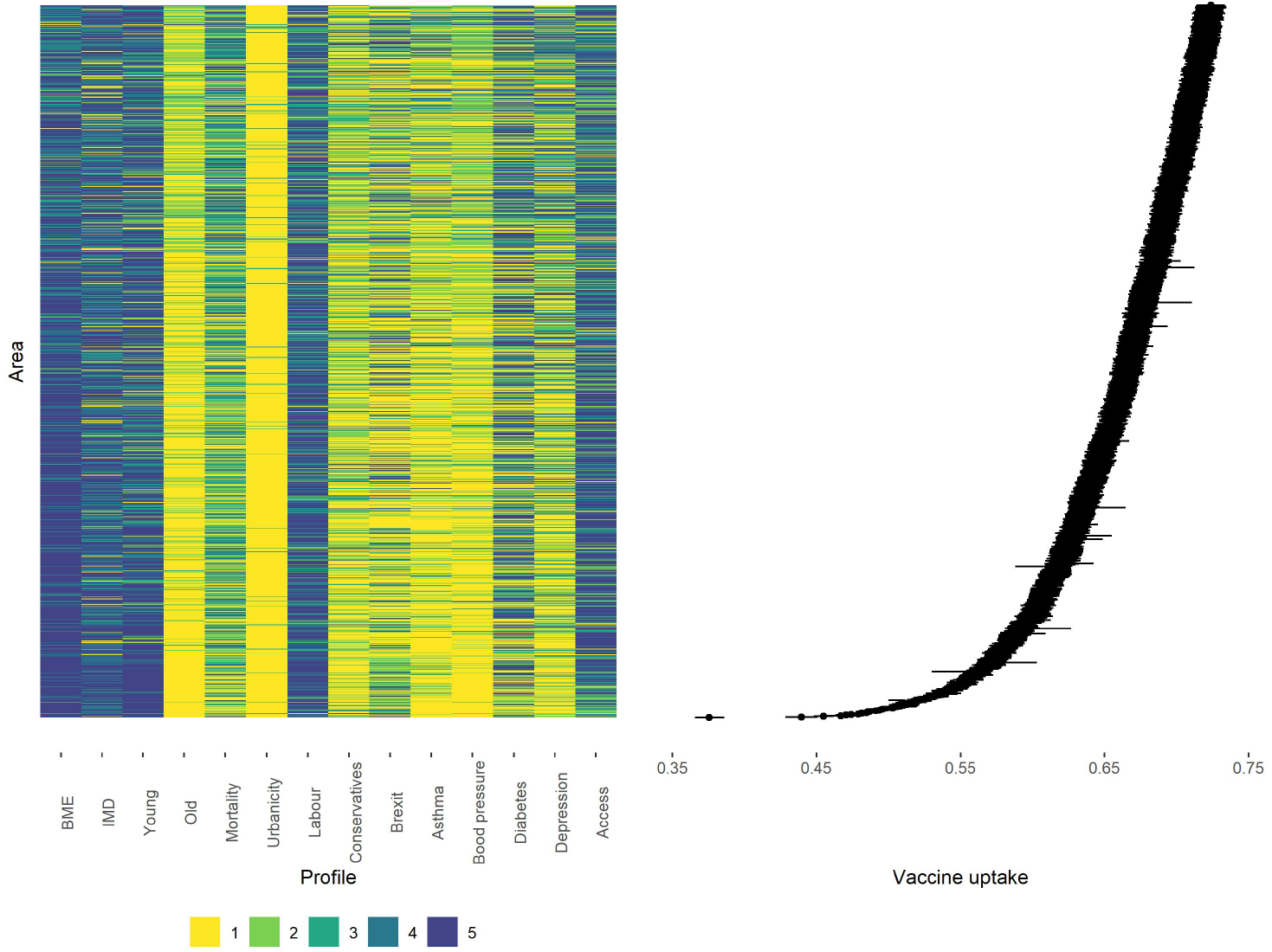
Covariate profiles for the areas characterised by the posterior median of vaccination uptake in the first quintile (left) and 95%CI vaccine uptake rates (right).

While Figure 2 provides a descriptive characterisation of the areas with low and high vaccination uptake, Figure 3 visualises the strength of the relationship between community-level characteristics and vaccination uptake in England, by means of OR and 95%CI. The index of multiple deprivation is the covariate most strongly associated with low vaccine coverage; the odds of being vaccinated when living in the most deprived areas are 0.55 (95%CI 0.54-0.57) times those in the least deprived ones. Similarly the most ethnically diverse areas (highest quintile of BME population) have a 38% (95%CI 36-40%) lower odds of vaccine uptake compared with the least ethnically diverse. Additionally, areas with a higher proportion of population between 12 and 24 years old had lower odds of vaccination uptake (OR 0.87, 95%CI 0.85-0.89). Areas voting for conservatives at the 2019 general elections have higher vaccine coverage (OR 1.09, 95%CI 1.04-1.14), while there is not enough evidence of an association for the proportion of people voting labour and for the Brexit referendum. There is also insufficient evidence of an association between urbanicity with COVID-19 vaccine uptake in the multivariable setting, despite a negative association seen in the univariate model, likely due to the correlation with the percentage of BME population, Figure S6 in Supplementary Material. Disease awareness and targeting of high risk groups, represented by the COVID-19 mortality rates and prevalence of pre-existing conditions show a relationship with vaccine coverage. In particular, areas having suffered the highest COVID-19 related mortality before the start of the vaccination campaign have higher odds (OR 1.08, 95%CI 1.05-1.11); similarly for areas with high prevalence of asthma or high blood pressure the ORs are 1.20, (95%CI 1.16-1.24) and 1.10 (95%CI 1.07-1.14) respectively. For diabetes and depression the link is less clear, and could potentially be affected by their correlation with the other health variables (Kendall’s tau is 0.33 between quintiles of diabetes and blood pressure and 0.43 between asthma and depression, see Figure S6 in Supplementary Material). Finally increase in vaccine accessibility improves COVID-19 vaccine coverage (OR 1.07, 95%CI 1.03-1.12).

**Fig. 3.**
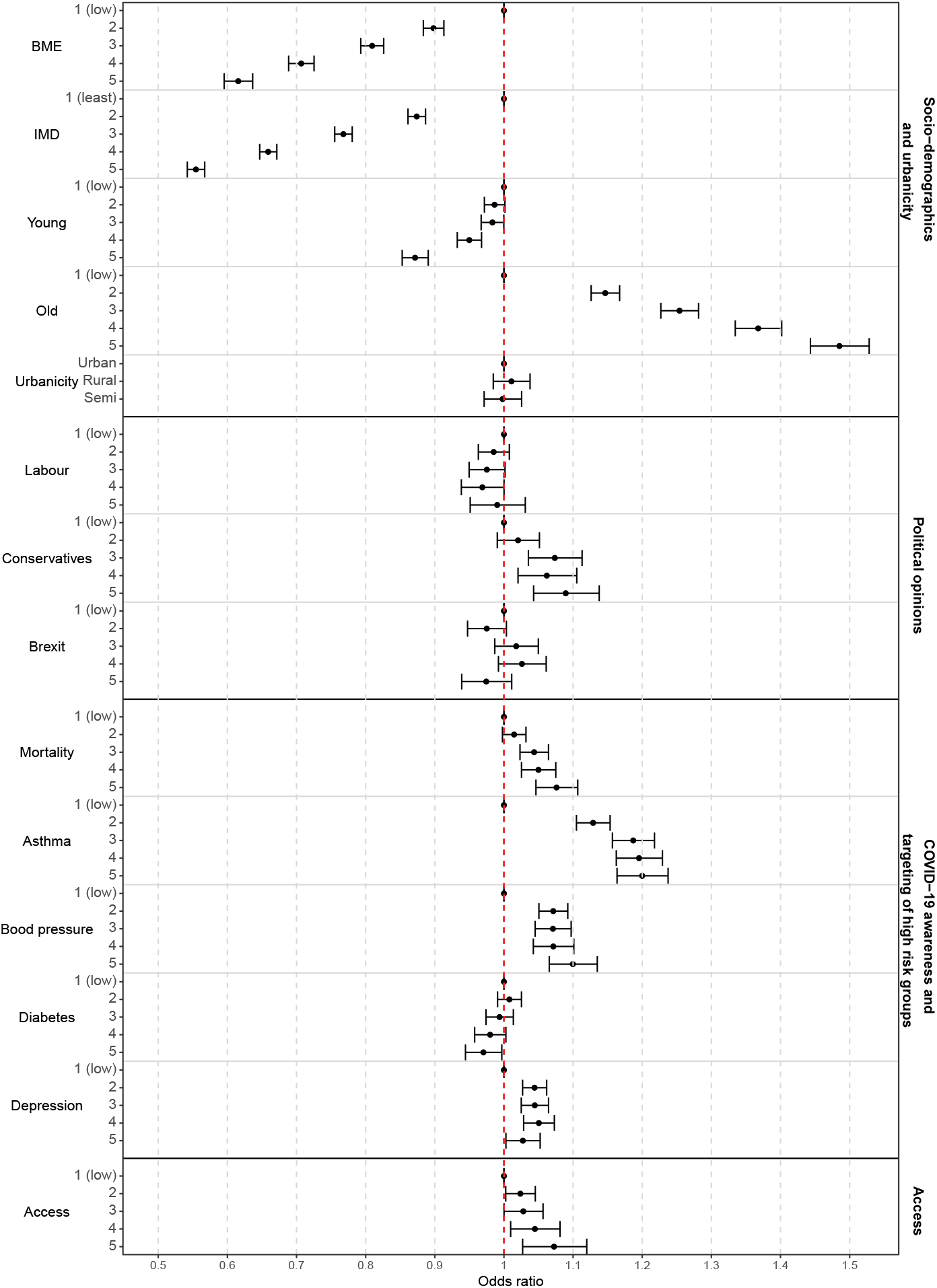
Posterior median odd ratios and 95%CI for the middle super output areas (MSOAs) characteristics and COVID-19 vaccination uptake.

The community-level characteristics included in the model explain 66% (95% CI 63%-69%) of the total variance of vaccination uptake. Figure 4 maps the posterior probability that the area-level odds of vaccination are lower than the national average, after accounting for the selected covariates. There is strong evidence (posterior probability higher than 0.8) that certain areas have lower odds of vaccination coverage due to unknown spatial covariates. The region around Manchester and Liverpool as well as the South East including London and part of the South West around Bristol show the highest probability of having lower odds (in yellow). Some level of geographical discrepancy is also visible in the 4 most populated cities in England, revealing unmeasured spatial confounding also in the large urban centers.

**Fig. 4.**
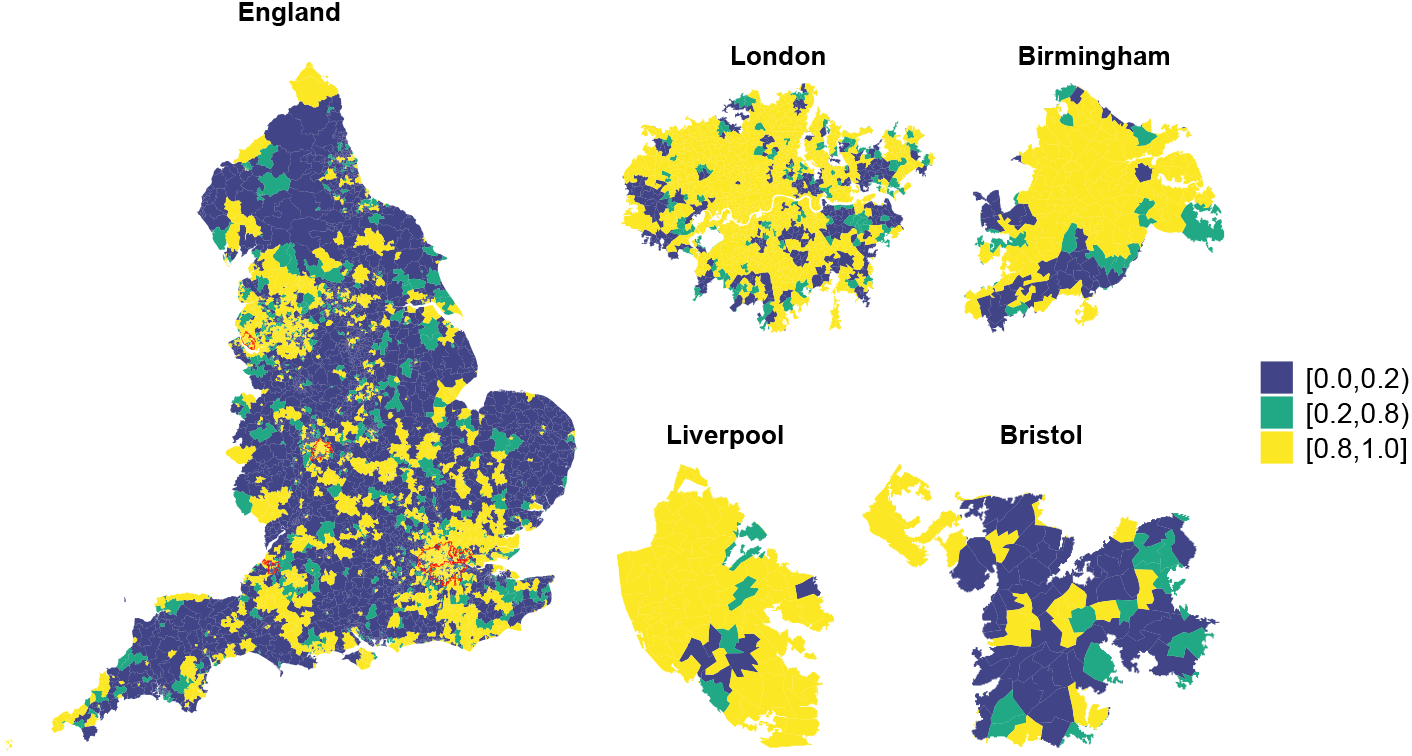
Posterior probability that the area-level odds of vaccination coverage are lower than the national average in England (left) and in the four largest cities (right).

## 4 Discussion

In this study we modelled the spatial variability of COVID-19 vaccine coverage in England at high spatial resolution a year after the mass vaccination started. We investigated the role of a range of community-level characteristics covering socio-demographics, awareness of COVID-19 health risks and targeting of high risk groups, political view and vaccine accessibility. Our model suggests that MSOAs with low COVID-19 vaccine uptake are the most deprived, ethnically diverse and with a higher proportion of young people. Awareness of COVID-19 health risk and accessibility to vaccine centres are also strongly associated with increased COVID-19 vaccine uptake. Areas where the majority of people voted for the Conservative party in the 2019 general election show higher uptake. Our findings add to previously reported evidence highlighting the role of age, ethnicity and socio-economic factors on vaccination refusal or delay, at the individual (e.g. [15, 48]) and aggregated level in England and internationally (e.g. [10, 49]). Additionally, in line with [50], who found that people with a physical health condition before the pandemic were more likely to take up the offer of vaccination, we also highlighted that areas more aware of the health risk due to COVID-19 are characterised by higher uptake. We also show that areas around the largest cities are more likely to be hesitant to the vaccination, in line with [51], a cross-sectional spatial modelling study in the UK conducted prior to the mass vaccination campaign, which revealed that London, Greater Manchester and Liverpool regions and minority ethnic groups were more resistant towards a new vaccine. Accessibility is found to have an impact on vaccination coverage, despite having a negative correlation with urbanicity. This is in line with [20] which showed that degree of urbanicity and population density had impact on vaccination site accessibility.

The main strength of our study is that it is the first to cover the entire population over 12 years old in England. As we focused on the first dose of the vaccine and considered the entire 2021, every individual over that age will have had the opportunity to receive the vaccine. In addition, as public health policies (nationally and internationally) regarding vaccination have changed and awareness was raised, people’s behaviour and attitude towards vaccination might have changed throughout 2021. Hence, using data covering an extended period makes our results generalisable and more relevant for population based public health policies. We considered a wide range of characteristics to capture vaccination inequalities and scepticism stemming from the different socio-demographic characteristics, political opinions, awareness of COVID-19 health risks and the targeting of high risk groups, and accessibility. Considering a high spatial resolution minimises ecological bias; at the same time the inclusion of spatial random effects ensures that we account for spatial variation due to unmeasured variables. As we showed in our results, there was still 30% unexplained variability in vaccination uptake, proving that it is necessary to consider spatial residuals in order to avoid potential biases in the inference. Our study has some limitations: some of the community-level covariates considered are not up-to-date; for instance information on BME population are related to the 2011 Census, hence we are implicitly assuming that the ethnic composition at MSOA level remains the same in the last decade. Furthermore, some variables are available at a coarser spatial resolution: COVID-19 mortality is at Local Authority districts (LAD), while political views are available at constituency level. In the analysis we assign the same value to all the MSOAs within the same LAD or constituency, leading to an underestimate of the variability at MSOA level and potentially a reduction in the association with those variables. Finally, the ecological nature of the study means that we cannot infer causal links between covariates and vaccination uptake [27].

Despite the good coverage of COVID-19 vaccine uptake at the national level, our results suggest that a year after the start of the vaccination campaign there are still substantial inequalities, most importantly related to deprivation and ethnicity. These have been highlighted from the beginning of the campaign (e.g. [14]) and have been later linked to general distrust in vaccines, low perception of risk as well as cultural/religious barriers [12, 52]. As they persist while the pandemic evolves, it is necessary to prioritise engagement through relevant figures, such as general practitioners, scientists and a wide spectrum of role models coming from these target communities [14].

The lower uptake in areas characterised by younger population should also be addressed. Several mechanisms have been investigated to encourage vaccinations among youngsters, focusing on highlighting the social benefit for the wider community [53]. As they generally perceive lower risk from the disease, it is important to stress the potential long-term impact. Additionally, education towards vaccination should provide clear messages and make use of trustworthy and similar messengers (for instance social media influencers,[54]) Accessibility remains a crucial underlying determinant for vaccination coverage and has a strong interplay with other variables: while the effort in expanding the vaccination centres network have substantially benefited the suburban and more rural areas in England [20], urban centers are still showing some of the lowest uptake, despite having a better access to vaccination points. As large cities are generally characterised by younger and more ethnically diverse populations it is crucial to tailor the access to vaccination to reach these subgroups, for instance using familiar locations such as schools, universities, community and language centres [52, 54].

Our results are also indicative of spatial variation in the vaccination coverage that could not be explained by the selected covariates. These spatial residuals are likely to capture variations that the community-level characteristics cannot adequately represent, either due to data availability (older data or at coarser geographical resolution) or definitions (for instance the definition of accessibility). In addition, the spatial residuals could reflect covariates we might have missed, such as occupation, which could capture part of the observed spatial discrepancies, as for instance health and social workers had more pressure to get vaccinated. Finally, the residual spatial trends could reflect area-level vaccine scepticism, which could be prevalent across various socioeconomic, cultural, ethnic and religious backgrounds [55]. This could also be the result of social exclusion, poor experiences at health services, false information, or a lack of trust in authorities and institutions [55].

To conclude, in our study we found that there are still marked geographical variations in vaccine uptake in England. We highlighted the role of community-level characteristics in explaining this variability, and showed how deprivation, ethnicity, age structure and accessibility are the most relevant. We also observed strong unknown spatial confounding which might reflects, at least in part, community-level vaccine scepticism. In order to level up the inequalities in vaccination uptake, actions are necessary to engage marginalised communities by implementing active outreach and using trusted sources such as general practitioners, scientists and influencers to respond to concerns about vaccine safety and efficacy.

## Data Availability

All data produced are available online at https://github.com/Georges3/COVID_19-VaccineUptake

## Supplementary information

Supplementary text, tables and Figures are available online.

## Acknowledgments

All authors acknowledge infrastructure support for the Department of Epidemiology and Biostatistics provided by the NIHR Imperial Biomedical Research Centre (BRC). We also thank prof. Paolo Vineis for insightful comments on the paper draft.

## Declarations

### Contributors

M.B: Conceptualisation, methodology, supervision, writing - original draft, writing - review and editing

G.K: Methodology, supervision, visualisation, writing - review and editing

G.B. Data preparation, statistical analyses, visualisation, writing - original draft.

All the authors read and approved the final draft.

### Patient consent for publication

Not required.

### Ethics

The study uses secondary, aggregated, anonymised data which are publicly available. No ethical permission was required.

### Data availability statement

The paper uses publicly available data at MSOA level. The final dataset used for the analyses and code for reproducing the results are available on the GitHub repository https://github.com/Georges3/COVID_19-VaccineUptake.

## References

[1] Wickaware, C.: Everything you need to know about the UK’s COVID-19 vaccination programme. Pharmaceutical Journal 306, 1424 (2021)

[2] The Institute for Government. https://www.instituteforgovernment.org.uk/printpdf/10196. Accessed: 2021-15-12

[3] Vaccinations in United Kingdom. https://coronavirus.data.gov.uk/details/vaccinations. Accessed: 2022-01-01

[4] Kadambari, S., Vanderslott, S.: Lessons about COVID-19 vaccine hesitancy among minority ethnic people in the UK. The Lancet Infectious Diseases 21(9), 1204–1206 (2021). https://doi.org/10.1016/S1473-3099(21)00404-7

[5] Burke, P.F., Masters, D., Massey, G.: Enablers and barriers to COVID-19 vaccine uptake: An international study of perceptions and intentions. Vaccine 39(36), 5116–5128 (2021). https://doi.org/10.1016/j.vaccine.2021.07.056

[6] Robinson, E., Jones, A., Daly, M.: International estimates of intended uptake and refusal of COVID-19 vaccines: A rapid systematic review and meta-analysis of large nationally representative samples. Vaccine 39(15), 2024–2034 (2021). https://doi.org/10.1016/j.vaccine.2021.02.005

[7] Duan, Y., Shi, J., Wang, Z., Zhou, S., Jin, Y., Zheng, Z.-J.: Disparities in COVID-19 vaccination among low-, middle-, and high-income countries: the mediating role of vaccination policy. Vaccines 9(8), 905 (2021). https://doi.org/10.3390/vaccines9080905

[8] Khaled, S.M., Petcu, C., Bader, L., Amro, I., Al-Hamadi, A.M.H., Al Assi, M., Ali, A.A.M., Le Trung, K., Diop, A., Bellaj, T., et al.: Prevalence and potential determinants of COVID-19 vaccine hesitancy and resistance in qatar: results from a nationally representative survey of qatari nationals and migrants between december 2020 and january 2021. Vaccines 9(5), 471 (2021). https://doi.org/10.3390/vaccines9050471

[9] Green, M.S., Abdullah, R., Vered, S., Nitzan, D.: A study of ethnic, gender and educational differences in attitudes toward COVID-19 vaccines in israel–implications for vaccination implementation policies. Israel Journal of Health Policy Research 10(1), 1–12 (2021). https://doi.org/10.1186/s13584-021-00458-w

[10] Mollalo, A., Tatar, M.: Spatial modeling of COVID-19 vaccine hesitancy in the united states. International Journal of Environmental Research and Public Health 18(18), 9488 (2021). https://doi.org/10.3390/ijerph18189488

[11] Shih, S.-F., Wagner, A.L., Masters, N.B., Prosser, L.A., Lu, Y., Zikmund-Fisher, B.J.: Vaccine hesitancy and rejection of a vaccine for the novel coronavirus in the united states. Frontiers in Immunology 12, 2275 (2021). https://doi.org/10.3389/fimmu.2021.558270

[12] Robertson, E., Reeve, K.S., Niedzwiedz, C.L., Moore, J., Blake, M., Green, M., Katikireddi, S.V., Benzeval, M.J.: Predictors of COVID-19 vaccine hesitancy in the UK household longitudinal study. Brain, Behavior, and Immunity 94, 41–50 (2021). https://doi.org/10.1016/j.bbi.2021.03.008

[13] Williams, L., Flowers, P., McLeod, J., Young, D., Rollins, L., et al.: Social patterning and stability of intention to accept a covid-19 vaccine in scotland: will those most at risk accept a vaccine? Vaccines 9(1), 17 (2021). https://doi.org/10.3390/vaccines9010017

[14] Perry, M., Akbari, A., Cottrell, S., Gravenor, M.B., Roberts, R., Lyons, R.A., Bedston, S., Torabi, F., Griffiths, L.: Inequalities in coverage of COVID-19 vaccination: A population register based cross-sectional study in Wales, UK. Vaccine 39(42), 6256–6261 (2021). https://doi.org/10.1016/j.vaccine.2021.09.019

[15] Gaughan, C.H., Razieh, C., Khunti, K., Banerjee, A., Chudasama, Y.V., Davies, M.J., Dolby, T., Gillies, C.L., Lawson, C., Mirkes, E.M., et al.: COVID-19 vaccination uptake amongst ethnic minority communities in england: a linked study exploring the drivers of differential vaccination rates. Journal of Public Health (Oxford, England) (2022). https://doi.org/10.1093/pubmed/fdab400

[16] Nafilyan, V., Dolby, T., Razieh, C., Gaughan, C.H., Morgan, J., Ayoubkhani, D., Walker, S., Khunti, K., Glickman, M., Yates, T.: Sociodemographic inequality in COVID-19 vaccination coverage among elderly adults in England: a national linked data study. BMJ open 11(7), 053402 (2021). https://doi.org/10.1136/bmjopen-2021-053402

[17] Coronavirus and vaccine hesitancy, Great Britain: 9 August 2021. https://www.ons.gov.uk/peoplepopulationandcommunity/healthandsocialcare/healthandwellbeing/bulletins/coronavirusandvaccinehesitancygreatbritain/9august2021. Accessed: 2021-18-12

[18] Viswanath, K., Bekalu, M., Dhawan, D., Pinnamaneni, R., Lang, J., McLoud, R.: Individual and social determinants of COVID-19 vaccine uptake. BMC Public Health 21(1), 1–10 (2021). https://doi.org/10.1186/s12889-021-10862-1

[19] Agarwal, R., Dugas, M., Ramaprasad, J., Luo, J., Li, G., Gao, G.G.: Socioeconomic privilege and political ideology are associated with racial disparity in covid-19 vaccination. Proceedings of the National Academy of Sciences 118(33) (2021). https://doi.org/10.1073/pnas.2107873118

[20] Duffy, C., Newing, A., Górska, J.: Evaluating the geographical accessibility and equity of COVID-19 vaccination sites in england. Vaccines 10(1), 50 (2022). https://doi.org/10.3390/vaccines10010050

[21] Cook, E.J., Elliott, E., Gaitan, A., Nduka, I., Cartwright, S., Egbutah, C., Randhawa, G., Waqar, M., Ali, N.: Vaccination against COVID-19: Factors that influence vaccine hesitancy among an ethnically diverse community in the UK. Vaccines 10(1), 106 (2022). https://doi.org/10.3390/vaccines10010106

[22] Office for National Statistics. Coronavirus and the social impacts on Great Britain, 29 Jan 2021. https://www.ons.gov.uk/peoplepopulationandcommunity/healthandsocialcare/healthandwellbeing/bulletins/coronavirusandthesocialimpactsongreatbritain/29january2021. Accessed: 2021-18-12

[23] Royal Society for Public Health. New poll finds BAME groups less likely to want COVID-19 vaccine. 2020. https://www.rsph.org.uk/about-us/news/new-poll-finds-bame-groups-less-likely-to-want-covid-vaccine.html. Accessed: 2022-03-01

[24] Freeman, D., Loe, B.S., Chadwick, A., Vaccari, C., Waite, F., Rosebrock, L., Jenner, L., Petit, A., Lewandowsky, S., Vanderslott, S., et al.: COVID-19 vaccine hesitancy in the UK: the oxford coronavirus explanations, attitudes, and narratives survey (oceans) II. Psychological Medicine, 1–15 (2020). https://doi.org/10.1017/S0033291720005188

[25] Razai, M.S., Osama, T., McKechnie, D.G., Majeed, A.: COVID-19 vaccine hesitancy among ethnic minority groups. British Medical Journal Publishing Group (2021). https://doi.org/10.1136/bmj.n513

[26] Scientific Advisory Group for Emergencies: Factors influencing COVID-19 vaccine uptake among minority ethnic groups. SAGE London (2020)

[27] Wakefield, J.: Ecologic studies revisited. Annual Reviews of Public Health 29, 75–90 (2008). https://doi.org/10.1146/annurev.publhealth.29.020907.090821

[28] Department of Health and Social Care, UK COVID-19 vaccines delivery plan, 11 January 2021. https://assets.publishing.service.gov.uk/government/uploads/system/uploads/attachment_data/file/951284/UK_COVID-19_vaccines_delivery_plan.pdf. Accessed: 2021-06-12

[29] Vaccination in United Kingdom. https://coronavirus.data.gov.uk/details/download. Accessed: 2022-05-01

[30] Middle Layer Super Output Area. Available online. https://datadictionary.nhs.uk/nhsbusiness_definitions/middle_layer_super_output_area.html. Accessed: 2021-11-12

[31] COVID-19 confirmed deaths in England (to 31 December 2020): report. https://www.england.nhs.uk/coronavirus/publication/vaccination-sites/. Accessed: 2022-11-02

[32] Middle Super Output Area boundaries 2011 England and Wales. https://data.cambridgeshireinsight.org.uk/dataset/output-areas/resource/0e5ac3b8-de71-4123-a334-0d1506a50288. Accessed: 2021-11-12

[33] Office for Health Improvement Disparities. Public Health Profiles. https://fingertips.phe.org.uk. Accessed: 2021-15-11

[34] Ministry of Housing, Communities and Local Government. https://data-communities.opendata.arcgis.com/datasets/5e1c399d787e48c0902e5fe4fc1ccfe3/about. Accessed: 2021-05-12

[35] Middle Super Output Area population estimates: Office for National Statistics. https://www.ons.gov.uk/peoplepopulationandcommunity/populationandmigration/populationestimates/datasets/middlesuperoutputareamidyearpopulationestimates. Accessed: 2022-03-01

[36] Death registrations and occurrences by local authority and place of death. https://www.ons.gov.uk/datasets/weekly-deaths-local-authority/ editions. Accessed: 2021-16-12

[37] Rural Urban Classification. https://www.gov.uk/government/statistics/statistical-digest-of-rural-england. Accessed: 2022-05-01

[38] House of Commons Library. https://commonslibrary.parliament.uk/research-briefings/cbp-8749/. Accessed: 2021-06-12

[39] House of Commons Library. https://commonslibrary.parliament.uk/brexit-national-identity-and-ethnicity-in-the-referendum/. Accessed: 2021-07-12

[40] Constituency data: Health conditions. https://commonslibrary.parliament.uk/constituency-data-how-healthy-is-your-area/. Accessed: 2022-05-01

[41] NHS England. List of Vaccination Sites—10 February 2022. https://www.england.nhs.uk/coronavirus/publication/vaccination-sites/. Accessed: 2022-11-02

[42] Middle Layer Super Output Areas Population Weighted Centroids. https://geoportal.statistics.gov.uk/datasets/ons::middle-layer-super-output-areas-december-2011-population-weighted-centroids/about. Accessed: 2022-10-02

[43] Wakefield, J.C., Best, N., Waller, L.: Bayesian approaches to disease mapping. Spatial Epidemiology: Methods and Applications, 104–127 (2000). https://doi.org/10.1093/acprof:oso/9780198515326.003.0007

[44] Best, N., Richardson, S., Thomson, A.: A comparison of bayesian spatial models for disease mapping. Statistical Methods in Medical Research 14(1), 35–59 (2005). https://doi.org/10.1191/0962280205sm388oa

[45] Simpson, D., Rue, H., Riebler, A., Martins, T.G., Sørbye, S.H.: Penalising model component complexity: A principled, practical approach to constructing priors. Statistical Science 32(1), 1–28 (2017). https://doi.org/10.1214/16-STS576

[46] Rue, H., Martino, S., Chopin, N.: Approximate bayesian inference for latent gaussian models by using integrated nested laplace approximations. Journal of the Royal Statistical Society: Series B 71(2), 319–392 (2009). https://doi.org/10.1111/j.1467-9868.2008.00700.x

[47] The 1000 largest cities and towns in the United Kingdom in order of population. https://www.thegeographist.com/uk-cities-population-1000/. Accessed: 2022-11-02

[48] Soares, P., Rocha, J.V., Moniz, M., Gama, A., Laires, P.A., Pedro, A.R., Dias, S., Leite, A., Nunes, C.: Factors associated with COVID-19 vaccine hesitancy. Vaccines 9(3), 300 (2021). https://doi.org/10.3390/vaccines9030300

[49] Chernyavskiy, P., Richardson, J.W., Ratcliffe, S.J.: COVID-19 vaccine uptake in united states counties: geospatial vaccination patterns and trajectories towards herd immunity. medRxiv (2021). https://doi.org/10.1101/2021.05.28.21257946

[50] Batty, G.D., Deary, I.J., Altschul, D.: Pre-pandemic mental and physical health as predictors of COVID-19 vaccine hesitancy: evidence from a UK-wide cohort study. Annals of Medicine 54(1), 274–282 (2022). https://doi.org/10.1080/07853890.2022.2027007

[51] de Figueiredo, A.: Sub-national forecasts of COVID-19 vaccine acceptance across the UK: a large-scale cross-sectional spatial modelling study. medRxiv, 2020–12 (2021). https://doi.org/10.1101/2020.12.17.20248382

[52] Factors influencing COVID-19 vaccine uptake among minority ethnic group. https://assets.publishing.service.gov.uk/government/uploads/system/uploads/attachmentdata/file/952716/s0979-factors-influencing-vaccine-uptake-minority-ethnic-groups.pdf. Accessed: 2022-02-21

[53] Bell, S., Clarke, R., Mounier-Jack, S., Walker, J.L., Paterson, P.: Parents’ and guardians’ views on the acceptability of a future COVID-19 vaccine: A multi-methods study in england. Vaccine 38(49), 7789–7798 (2020). https://doi.org/10.1016/j.vaccine.2020.10.027

[54] Encouraging vaccine take-up among younger people. https://assets.publishing.service.gov.uk/government/uploads/system/uploads/attachmentdata/file/952716/s0979-factors-influencing-vaccine-uptake-minority-ethnic-groups.pdf.Accessed: 2022-02-21

[55] Simas, C., Larson, H.J.: Overcoming vaccine hesitancy in low-income and middle-income regions. Nature Reviews Disease Primers 7(1), 1–2 (2021). https://doi.org/10.1038/s41572-021-00279-w

