## Supplementary material for "Community-level characteristics of COVID-19 vaccine hesitancy in England: A nationwide cross-sectional study": -

Bucyibaruta Georges[1], Blangiardo Marta[1*]
and Konstantinoudis Garyfallos[1]

[1]MRC Centre for Environment and Health, Department of Epidemiology and Biostatistics, School of Public Health, Imperial College London, London, UK.

*Corresponding author(s). E-mail(s):
;
Contributing authors:;
;

### S1 Vaccine accessibility

In this section, we provide details on how we derived the variable that represents vaccination accessibility. By the end of February 2022, there were 2860 vaccination sites across England,with 99% of the country living within ten miles of a site. The sites are classified into 4 categories: pharmacies, General Practitioner-led, vaccinations centers and hospital hubs. To obtain accessibility at MSOA we combine information of vaccination sites and MSOA centroids coordinates, using the following steps:

- Extract the coordinates of the vaccination sites.
- Extract the population weighted MSOA centroid coordinates
- Calculate the distance between each centroid and all the vaccination sites
- Define the accessible vaccination sites (AVS) as the number of sites within 10 miles of each MSOA centroid
- Access is then calculated as AVS divided by the eligible population ($\geq$ 12 years old) in each MSOA, then multiplied by 10,000:

$$Access = \frac{AVS}{Pop} * 10000$$

### S2 Prior distributions

By default, a Gaussian prior with mean and precision equal to zero is considered for the intercept. On the regression coefficients we specified Gaussian prior with zero mean and precision equal to 0.001. Priors for the parameters of the random effect (hyperparameters) are assigned by using the Penalized Complexity (PC) framework [1]. These are priors that penalize departure from a base model (assumes to have a constant relative risk over all areas, i.e. no spatial variation or equivalently infinite precision). In addition, these priors are defined using probability statements about the parameter on the appropriate scale. In this setting, the PC-prior for the precision $\tau$ is defined on the standard deviation $\sigma = \frac{1}{\sqrt{\tau}}$ as $P(\frac{1}{\sqrt{\tau}} > U) = \alpha$. It was recognised that the interpretation of $\frac{1}{\sqrt{\tau}}$ is not obvious; but in the re-parametrized Besag-York-Molliè model used in our specification (BYM2) $\tau$ represents the marginal precision, making it easier to relate it to the total residual relative risk. The parameter $\phi$ represents a fraction of the total variance which can be attributed to spatial dependency structure [1, 2]. Its PC-prior specification, using a probability statement, is defined a $P(\phi < U) = \alpha$. Finally, the parameters related to $\tau$ are $U = 1$ and $\alpha = 0.01$, while those related to $\phi$ are set as $U = 0.5$ and $\alpha = 0.5$, using the default values presented in [1, 2].

### Figures

Fig. S1: Distribution of the vaccinations sites by type (left) and of the Accessibility variable used in the model (right).

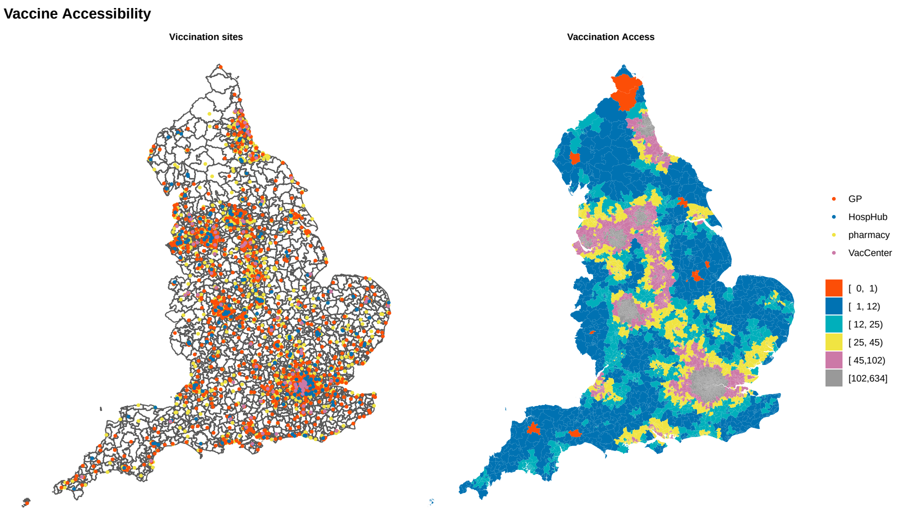

Fig. S2: Socio-Demographics and urbanicity related covariates

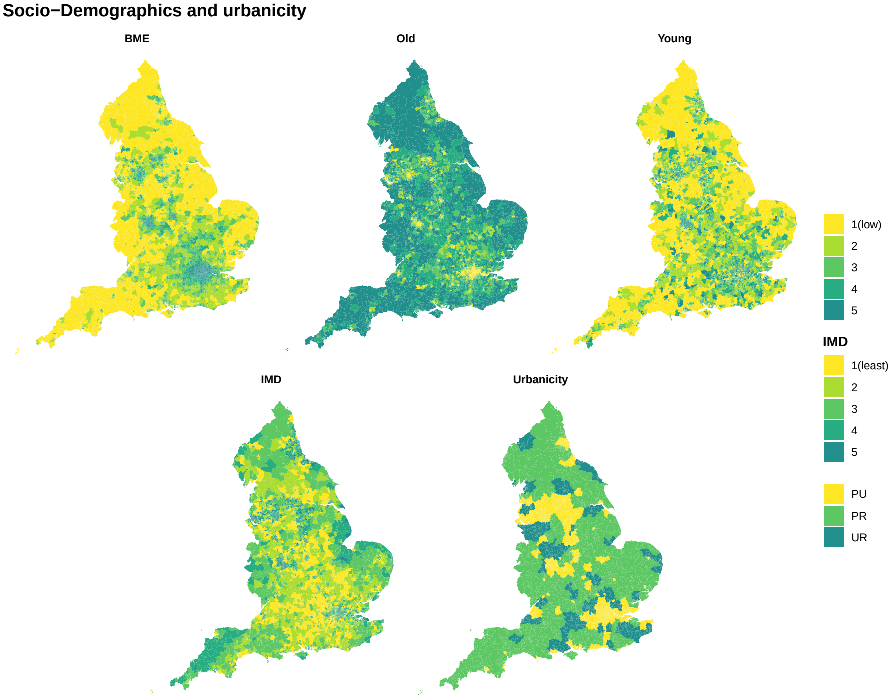

### Fig. S3: Political opinions and vaccine accessibility related covariates

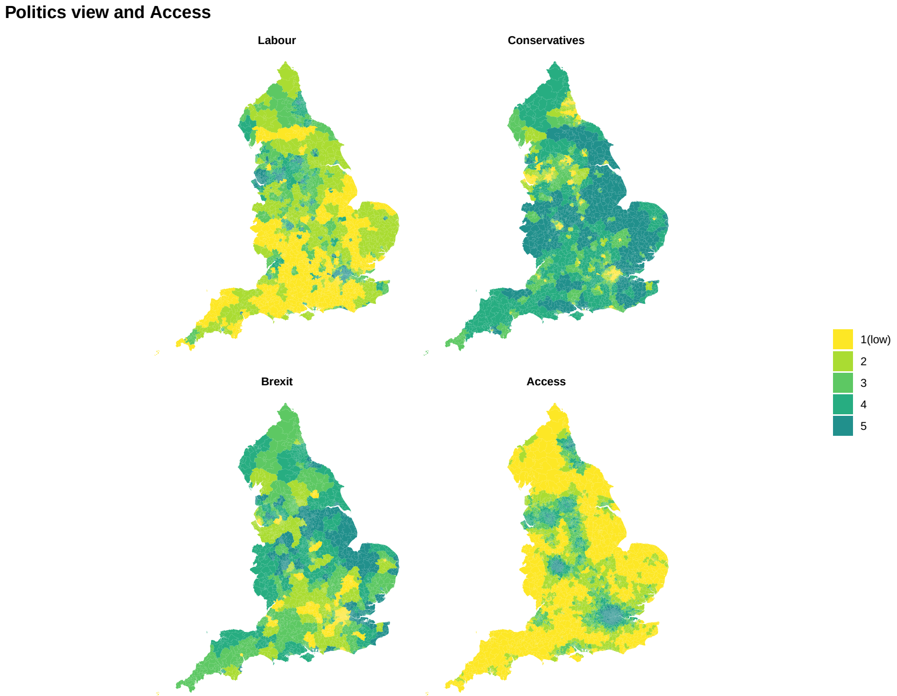

Fig. S4: COVID-19 awareness and targeting of high risk groups

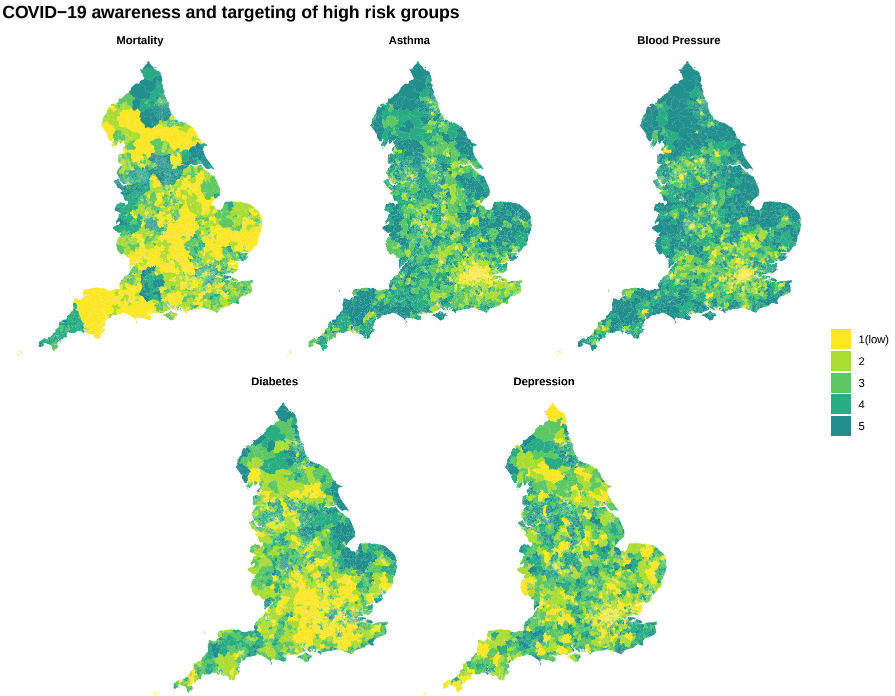

Fig. S5: Profiles of all covariates (left) and high vaccine uptake rates (right).

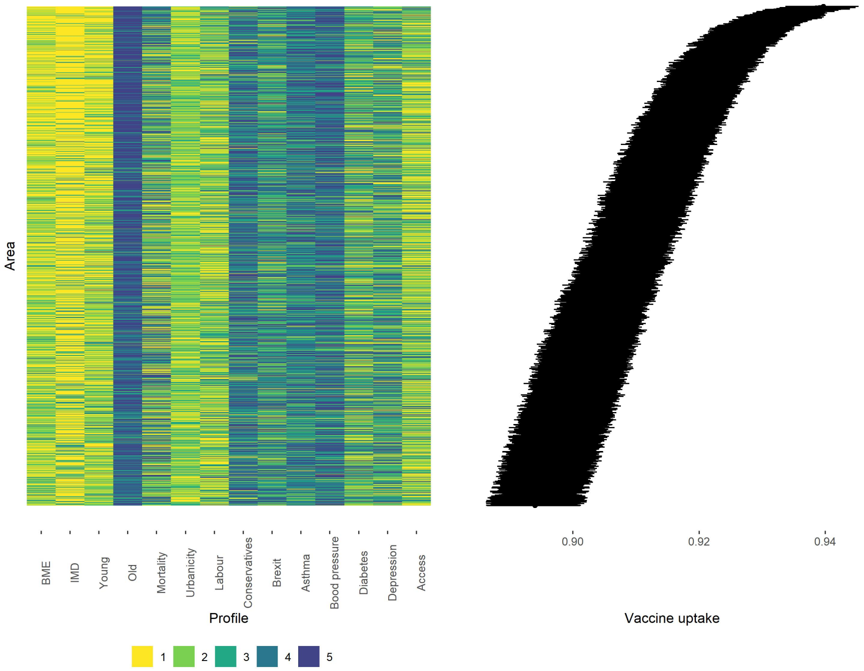

#### Fig. S6: Correlation Heatmap

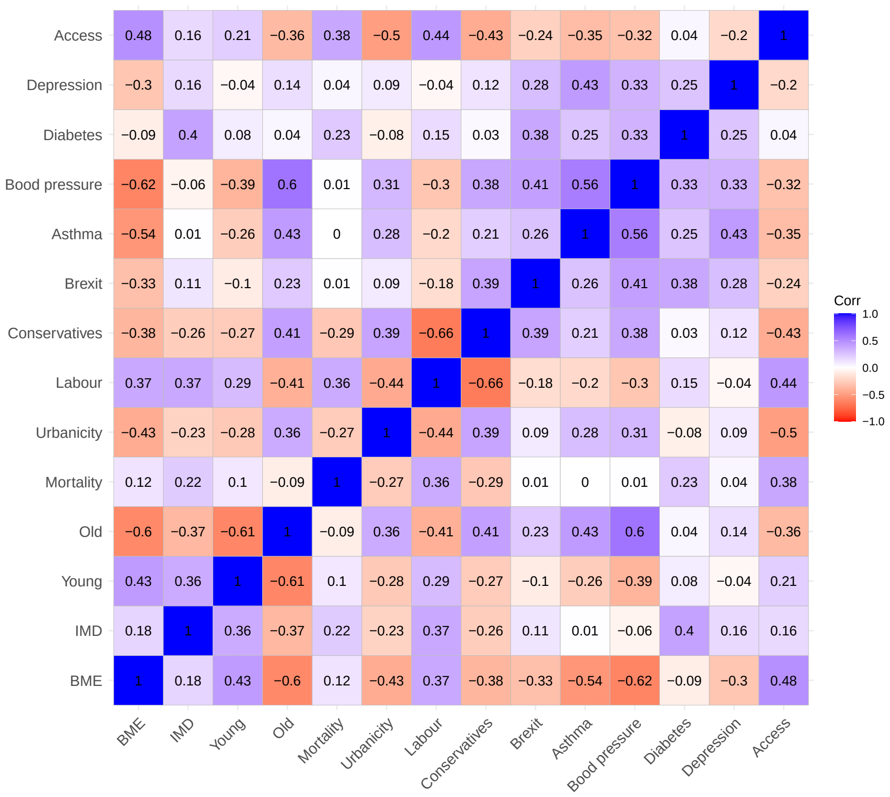

#### Fig. S7: Relationship between community-level characteristics and COVID-19 vaccination uptake in England (univariate models)

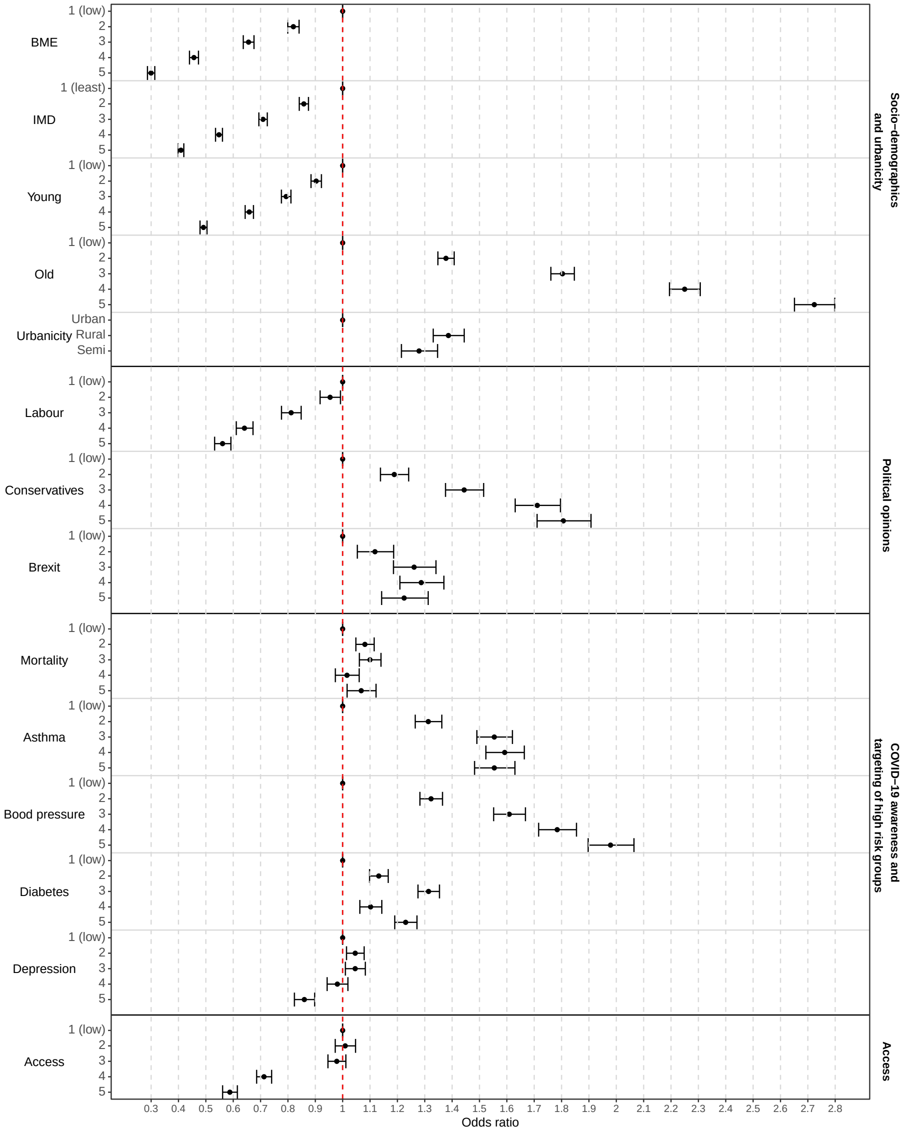

### Tables

Table S1: Quintiles of covariates considered in the analysis

| Variable | Min | Q1 | Q2 | Q3 | Q4 | Max |
|---|---|---|---|---|---|---|
| BME | 0.43 | 2.10 | 3.72 | 8.28 | 22.17 | 94.38 |
| IMD | 1.52 | 9.48 | 14.28 | 21.00 | 31.27 | 88.02 |
| Young | 6.48 | 14.09 | 15.49 | 16.87 | 19.14 | 83.46 |
| Old | 0.42 | 14.35 | 19.09 | 22.99 | 27.26 | 55.05 |
| Labour | 0 | 18.28 | 26.94 | 39.54 | 50.77 | 84.68 |
| Conservatives | 0 | 31.74 | 45.48 | 54.42 | 60.57 | 76.72 |
| Brexit | 21.38 | 46.03 | 51.69 | 56.52 | 62.05 | 75.56 |
| Access | 0 | 15.34 | 34.43 | 66.26 | 156.25 | 634.27 |
| Mortality | 0 | 0.18 | 0.23 | 0.28 | 0.34 | 0.65 |
| Asthma | 2.2 | 5.7 | 6.5 | 7.0 | 7.5 | 10.7 |
| Blood pressure | 1.8 | 12.2 | 14.0 | 15.3 | 16.8 | 27.2 |
| Diabetes | 1.2 | 6.0 | 6.8 | 7.5 | 8.3 | 15.0 |
| Depression | 3.4 | 9.1 | 10.8 | 12.4 | 14.4 | 27.0 |

Table S2: Odds ratio and 95%CI representing the relationship between community-level characteristics (Socio-demographics and urbanicity) and COVID-19 vaccination uptake in the multivariable model.

| | | mean | 2.5% | 97.5% |
|---|---|---|---|---|
| BME | 1 (low) | ref | | |
| | 2 | 0.90 | 0.88 | 0.91 |
| | 3 | 0.81 | 0.79 | 0.83 |
| | 4 | 0.71 | 0.69 | 0.72 |
| | 5 | 0.62 | 0.59 | 0.64 |
| IMD | 1 (least) | ref | | |
| | 2 | 0.87 | 0.86 | 0.89 |
| | 3 | 0.77 | 0.76 | 0.78 |
| | 4 | 0.66 | 0.65 | 0.67 |
| | 5 | 0.55 | 0.54 | 0.57 |
| Young | 1 (low) | ref | | |
| | 2 | 0.99 | 0.97 | 1.00 |
| | 3 | 0.98 | 0.97 | 1.00 |
| | 4 | 0.95 | 0.93 | 0.97 |
| | 5 | 0.87 | 0.85 | 0.89 |
| Old | 1 (low) | ref | | |
| | 2 | 1.15 | 1.13 | 1.17 |
| | 3 | 1.25 | 1.23 | 1.28 |
| | 4 | 1.37 | 1.33 | 1.40 |
| | 5 | 1.49 | 1.44 | 1.53 |
| Urbanicity | Urban | ref | | |
| | Rural | 1.01 | 0.98 | 1.04 |
| | Semi | 1.00 | 0.97 | 1.03 |

Table S3: Odds ratio and 95%CI representing the relationship between community-level characteristics (Political opinions and Vaccine accessibility) and COVID-19 vaccination uptake in the multivariable model.

| | | mean | 2.5% | 97.5% |
|---|---|---|---|---|
| Labour | 1 (low) | ref | | |
| | 2 | 0.98 | 0.96 | 1.01 |
| | 3 | 0.97 | 0.95 | 1.00 |
| | 4 | 0.97 | 0.94 | 1.00 |
| | 5 | 0.99 | 0.95 | 1.03 |
| Conservatives | 1 (low) | ref | | |
| | 2 | 1.02 | 0.99 | 1.05 |
| | 3 | 1.07 | 1.03 | 1.11 |
| | 4 | 1.06 | 1.02 | 1.10 |
| | 5 | 1.09 | 1.04 | 1.14 |
| Brexit | 1 (low) | ref | | |
| | 2 | 0.97 | 0.95 | 1.00 |
| | 3 | 1.02 | 0.99 | 1.05 |
| | 4 | 1.03 | 0.99 | 1.06 |
| | 5 | 0.97 | 0.94 | 1.01 |
| Access | 1 (low) | ref | | |
| | 2 | 1.02 | 1.00 | 1.04 |
| | 3 | 1.03 | 1.00 | 1.06 |
| | 4 | 1.04 | 1.01 | 1.08 |
| | 5 | 1.07 | 1.03 | 1.12 |

Table S4: Odds ratio and 95%CI representing the relationship between community-level characteristics (COVID-19 awareness and targeting of high risk groups) and COVID-19 vaccination uptake in the multivariable model.

| | | mean | 2.5% | 97.5% |
|---|---|---|---|---|
| Mortality | 1 (low) | ref | | |
| | 2 | 1.01 | 1.00 | 1.03 |
| | 3 | 1.04 | 1.02 | 1.06 |
| | 4 | 1.05 | 1.02 | 1.07 |
| | 5 | 1.08 | 1.05 | 1.11 |
| Asthma | 1 (low) | ref | | |
| | 2 | 1.13 | 1.10 | 1.15 |
| | 3 | 1.19 | 1.16 | 1.22 |
| | 4 | 1.20 | 1.16 | 1.23 |
| | 5 | 1.20 | 1.16 | 1.24 |
| Blood pressure | 1 (low) | ref | | |
| | 2 | 1.07 | 1.05 | 1.09 |
| | 3 | 1.07 | 1.04 | 1.10 |
| | 4 | 1.07 | 1.04 | 1.10 |
| | 5 | 1.10 | 1.07 | 1.14 |
| Diabetes | 1 (low) | ref | | |
| | 2 | 1.01 | 0.99 | 1.02 |
| | 3 | 0.99 | 0.97 | 1.01 |
| | 4 | 0.98 | 0.96 | 1.00 |
| | 5 | 0.97 | 0.94 | 1.00 |
| Depression | 1 (low) | ref | | |
| | 2 | 1.04 | 1.03 | 1.06 |
| | 3 | 1.04 | 1.02 | 1.06 |
| | 4 | 1.05 | 1.03 | 1.07 |
| | 5 | 1.03 | 1.00 | 1.05 |

Table S5: Odds ratio and 95%CI representing the relationship between community-level characteristics (Socio-demographics and urbanicity) and COVID-19 vaccination uptake in the univariate model.

| | | mean | 2.5% | 97.5% |
|---|---|---|---|---|
| BME | 1 (low) | ref | | |
| | 2 | 0.82 | 0.80 | 0.84 |
| | 3 | 0.66 | 0.64 | 0.68 |
| | 4 | 0.46 | 0.44 | 0.47 |
| | 5 | 0.30 | 0.29 | 0.31 |
| IMD | 1 (least) | 1.00 | 1.00 | 1.00 |
| | 2 | 0.86 | 0.84 | 0.88 |
| | 3 | 0.71 | 0.69 | 0.72 |
| | 4 | 0.55 | 0.54 | 0.56 |
| | 5 | 0.41 | 0.40 | 0.42 |
| Young | 1 (low) | ref | | |
| | 2 | 0.90 | 0.88 | 0.92 |
| | 3 | 0.79 | 0.78 | 0.81 |
| | 4 | 0.66 | 0.64 | 0.67 |
| | 5 | 0.49 | 0.48 | 0.50 |
| Old | 1 (low) | ref | | |
| | 2 | 1.38 | 1.35 | 1.41 |
| | 3 | 1.80 | 1.76 | 1.85 |
| | 4 | 2.25 | 2.19 | 2.31 |
| | 5 | 2.72 | 2.65 | 2.80 |
| Urbanicity | Urban | ref | | |
| | Rural | 1.39 | 1.33 | 1.44 |
| | Semi | 1.28 | 1.21 | 1.35 |

Table S6: Odds ratio and 95%CI representing the relationship between community-level characteristics (Political opinions and Vaccine accessibility) and COVID-19 vaccination uptake in the univariate model.

| | | mean | 2.5% | 97.5% |
|---|---|---|---|---|
| Labour | 1 (low) | ref | | |
| | 2 | 0.95 | 0.92 | 0.99 |
| | 3 | 0.81 | 0.78 | 0.85 |
| | 4 | 0.64 | 0.61 | 0.67 |
| | 5 | 0.56 | 0.53 | 0.59 |
| Conservatives | 1 (low) | ref | | |
| | 2 | 1.19 | 1.14 | 1.24 |
| | 3 | 1.44 | 1.38 | 1.52 |
| | 4 | 1.71 | 1.63 | 1.80 |
| | 5 | 1.81 | 1.71 | 1.91 |
| Brexit | 1 (low) | ref | | |
| | 2 | 1.12 | 1.05 | 1.19 |
| | 3 | 1.26 | 1.19 | 1.34 |
| | 4 | 1.29 | 1.21 | 1.37 |
| | 5 | 1.22 | 1.14 | 1.31 |
| Access | 1 (low) | ref | | |
| | 2 | 1.08 | 1.05 | 1.12 |
| | 3 | 1.10 | 1.06 | 1.14 |
| | 4 | 1.02 | 0.97 | 1.06 |
| | 5 | 1.07 | 1.02 | 1.12 |

Table S7: Odds ratio and 95%CI representing the relationship between community-level characteristics (COVID-19 awareness and targeting of high risk groups) and COVID-19 vaccination uptake in the univariate model.

| | | mean | 2.5% | 97.5% |
|---|---|---|---|---|
| Mortality | 1 (low) | 1.00 | 1.00 | 1.00 |
| | 2 | 1.31 | 1.27 | 1.36 |
| | 3 | 1.55 | 1.49 | 1.62 |
| | 4 | 1.59 | 1.52 | 1.66 |
| | 5 | 1.55 | 1.48 | 1.63 |
| Asthma | 1 (low) | 1.00 | 1.00 | 1.00 |
| | 2 | 1.32 | 1.28 | 1.36 |
| | 3 | 1.61 | 1.55 | 1.67 |
| | 4 | 1.78 | 1.72 | 1.85 |
| | 5 | 1.98 | 1.90 | 2.06 |
| Blood pressure | 1 (low) | 1.00 | 1.00 | 1.00 |
| | 2 | 1.13 | 1.10 | 1.17 |
| | 3 | 1.31 | 1.28 | 1.35 |
| | 4 | 1.10 | 1.06 | 1.14 |
| | 5 | 1.23 | 1.19 | 1.27 |
| Diabetes | 1 (low) | 1.00 | 1.00 | 1.00 |
| | 2 | 1.05 | 1.01 | 1.08 |
| | 3 | 1.05 | 1.01 | 1.08 |
| | 4 | 0.98 | 0.94 | 1.02 |
| | 5 | 0.86 | 0.82 | 0.90 |
| Depression | 1 (low) | 1.00 | 1.00 | 1.00 |
| | 2 | 1.01 | 0.97 | 1.05 |
| | 3 | 0.98 | 0.95 | 1.01 |
| | 4 | 0.71 | 0.69 | 0.74 |
| | 5 | 0.59 | 0.56 | 0.62 |

### References

[1] Riebler, A., Sørbye, S.H., Simpson, D., Rue, H.: An intuitive bayesian spatial model for disease mapping that accounts for scaling. Statistical methods in medical research **25**(4), 1145–1165 (2016). https://doi.org/10.1177/0962280216660421

[2] Simpson, D., Rue, H., Riebler, A., Martins, T.G., Sørbye, S.H.: Penalising model component complexity: A principled, practical approach to constructing priors. Statistical Science **32**(1), 1–28 (2017). https://doi.org/10.1214/16-STS576
